# Ibrutinib as a Potential Therapeutic for Cocaine Use Disorder

**DOI:** 10.1101/2021.02.05.21251228

**Authors:** Spencer B. Huggett, Jeffrey S. Hatfield, Joshua D. Walters, John E. McGeary, Justine W. Welsh, Trudy F. C. Mackay, Robert R. H. Anholt, Rohan H.C. Palmer

## Abstract

Cocaine use presents a worldwide public health problem with high socioeconomic cost. Current treatments for cocaine use disorder (CUD) are suboptimal and rely primarily on behavioral interventions. To explore pharmaceutical treatments for CUD, we analyzed genome-wide gene expression data from publically availble human brain tissues (midbrain, hippocampus and prefrontal cortex neurons) from 71 individuals (mean age = 39.9, 100% male, 36 with CUD and 35 matched controls). We leveraged the L1000 database to investigate molecular associations between neuronal mRNA profiles from 825 repurposable compounds (e.g., FDA approved) with human CUD gene expression in the brain. We identified 16 compounds that were negatively associated with CUD gene expression patterns across all brain regions (*p*_adj_ < 0.05), all of which outperformed current targets undergoing clinical trials for CUD (all *p*_adj_ > 0.05). We tested the effectiveness of these compounds using independent transcriptome-wide *in vitro* (neuronal cocaine exposure; n=18) and *in vivo* (mouse cocaine self-administration; prefrontal cortex, hippocampus and midbrain; n = 12-15) datasets. Among these medications, Ibrutinib demonstrated negative associations with both neuronal cocaine exposure and mouse cocaine self-administration. To obtain experimental confirmation of therapeutic effects of Ibrutinib on CUD, we used the *Drosophila melanogaster* model, which enables highthroughput quantification of behavioral responses in defined genetic backgrounds and controlled environmental conditions. Ibrutinib altered cocaine-induced changes in startle response and reduced the occurrence of cocaine-induced seizures (n = 61-142 per group; sex: 51%female). Our results identify Ibrutinib, an FDA approved medication, as a potential therapeutic for cocaine neurotoxicity.

## INTRODUCTION

Approximately 18.1 million people use cocaine globally each year^1^ and roughly 30% of those users reside in the United States^2^ (US). In the US, nearly 1 million individuals meet criteria for cocaine use disorder^2^ (CUD) and 13% of the individuals in North American substance use treatment facilities are treated for CUD^1^. Cocaine use increases risk for cardiovascular disease, seizures, and mental health disorders and contributes to roughly 40% of drug-related emergency room visits in the US^3^. Cocaine toxicity occurs via multiple mechanisms in nearly all tissue types^4^. Since 1999, cocaine-involved overdose deaths have increased by 416% in the US and resulted in 15,883 deaths in 2019^5^. To date, there are no medications approved by the Food and Drug Administration (FDA) for human CUD or cocaine toxicity.

Developing treatments for substance use disorders is a key priority of the National Institute on Drug Abuse. While behavioral treatments for CUD exist, they have limited efficacy and are plagued by high dropout rates^6^. Over 20 pharmaceutical medications are currently being tested for clinical trials for CUD. Most of these medications include antidepressants, antipsychotics, psychostimulants, cognitive enhancing drugs, anxiolytics, repurposed medications for other substance use disorders, anticonvulsants/muscle relaxants, and dopamine agonists^7^. These treatments generally have mixed efficacy and often only demonstrate effects in particular subpopulations. Medications for cocaine toxicity are also limited. Emergency room practioners rely on beta-blockers for cocaine-cardiotoxicity^8^ and benzodiazapines for cocaine neurotoxicity^9^ (e.g., seizures). Many candidate compounds for treating CUD and cocaine toxicity rely on repurposing existing medications, leveraging knowledge of cocaine’s pharmacology, and/or the biological mechanisms underpinning cocaine pathology.

The molecular neuropathology of CUD is characterized by persistent cellular and molecular adaptations across multiple brain regions, particularly in the mesocorticolimbic “reward” pathway. Molecular neuroadaptations in this pathway mediate reward, motivation, behavioral control, memory formation, incentive salience, cue, drug and stress-induced drug taking/relapse. One approach to investigate the neuromolecular correlates underlying CUD (and potentially cocaine neurotoxicity) is to examine gene expression profiles in post-mortem brain tissue by comparing individuals with CUD to matched cocaine-free controls. Currently, three studies are publicly available that assess CUD gene expression profiles in key addiction brain regions: the dorsal-lateral prefrontal cortex^10^ (dlPFC), hippocampus^11^ and midbrain^12^. Previously, medications that consistently target gene expression across meso-cortico-limbic brain regions have demonstrated robust effects for reducing binge drinking in mice^13^. Thus, screening medications that target transcriptional patterns across relevant brain regions may help identify FDA-approved therapeutics that could be repurposed for treating CUD.

Drug discovery analyses can cut costs and time for developing effective therapeutics, which on average cost $1.4 billion and take 12-16 years to develop^14^. The NIH-funded Library of Integrated Network-based Cellular Signatures (LINCS L1000) database facilitates drug development strategies by indexing over 1.3 million expression profiles resulting from an extensive catalog of more than 48,000 perturbagens (pharmaceuticals, small molecules, shRNAs, cDNAs, and biologics) in over 80 human cell lines^15^. This resource allows rapid screening of compounds that may target the molecular mechanisms of a disease.

Here, we outline a neuronal mRNA drug-repurposing framework that identifies and validates potential medications for CUD and/or cocaine neurotoxicity (see **Figure 1** for study overview).

**Figure 1:**
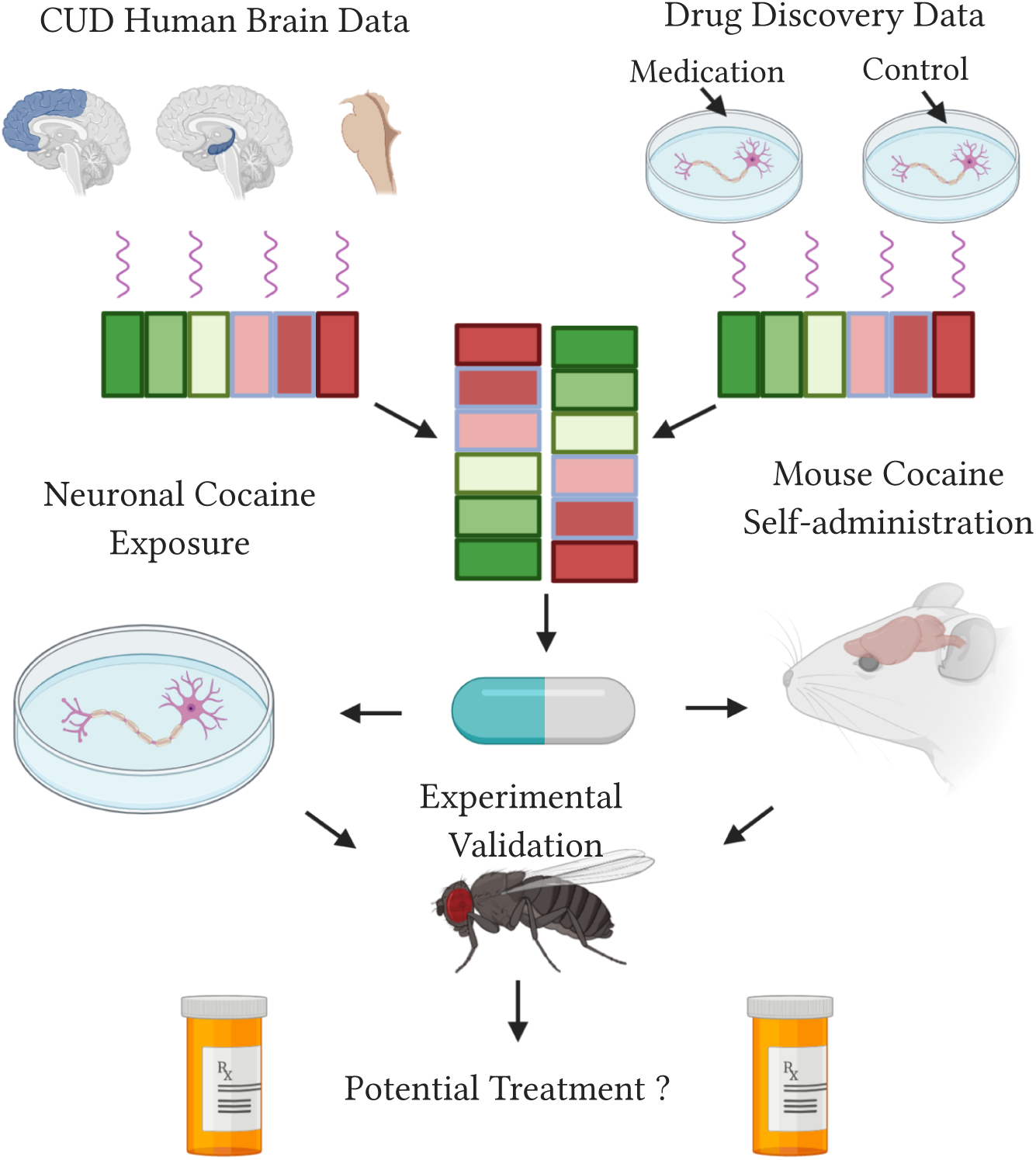
Overview of the study.

## METHODS

### Drug Discovery Input

Our human sample (n = 71) comprised of publicly available gene expression data from post-mortem brain tissue of individuals with cocaine use disorder (CUD; n = 36) and matched cocaine-free controls (n = 35). Data were obtained from three independent studies (GSM2642566^10^; SRA029279^11^; E-GEOD-54839^12^; Supplementary Table S1). We conducted drug discovery analyses leveraging the L1000 database, which catalogues mRNA expression profiles of human cell types exposed to therapeutic compounds^15^.

The input for our drug discovery analysis included three categories. The first category included all differentially expressed genes (Benjamini-Hochberg False Discovery Rate (FDR) < 0.05) associated with CUD in the dlPFC^16^, hippocampus^17^ and midbrain^12^. Differentially expressed genes might include genes attributed to cocaine-induced toxicity, cardiac complications or psychiatric comorbidities. Thus, our second category focused on the genes related to the behavioral manifestations of CUD, by including the mRNA expression of genes from the cocaine addiction pathway (Kyoto Encyclopedia of Genes and Genomes [KEGG] database). The last category sought to further minimize potential biases of measurement error by employing “landmark” genes (those directly measured from L1000, rather than imputed) that were part of at least one of the first two categories. We refer to the input genes for our drug discovery analysis as CUD genes (Supplementary File S1).

### Drug Discovery Analyses

Drug discovery analyses utilized the L1000 Connectivity Mapping gene expression profiles from the Library of Integrated Network Based Cellular Signatures (level 5 data from phase I: GSE92742 and phase II: GSE70138). Since the focus of the study was to find repurposable treatments for CUD, we selected all compounds that were FDA approved *or* in stages 1-3 of clinical trials – as listed from the drug repurposing hub website (https://clue.io/repurposing#download-data; updated 3/24/2020). We benchmarked our findings with treatments that were currently undergoing clinical trials for CUD and specifically those that were reported on the https://clinicaltrials.gov/website.

The mechanisms of action for most pharmacological treatments of substance use disorders target neuronal processes. Thus, we focused our drug discovery analysis on two neuronal cell types used in the L1000 database: differentiated neuronal cells and neuronal progenitor cells. In total, we evaluated the potential therapeutic value of 825 compounds, which spanned 3,468 individual neuronal mRNA signatures (*in vitro* gene expression profiles for a compound measured at a particular dose, time, and cell line). For each signature, we estimated a linear Pearson Product-Moment correlation coefficient with the CUD input. Note that not all medications undergoing clinical trials for CUD were included in the human neuronal cell lines from the L1000 database (Supplementary File S2).

To identify potential treatments for CUD, we conducted multi-level meta-regressions for all 825 compounds using the metafor package in R^18^. Multi-level meta-regression provides a powerful and interpretable framework that can accommodate for complex data-types. Our meta-regressions adjusted for two random effects: human brain region (or study) and the three input categories within studies (e.g. KEGG_dlPFC_, KEGG_Hippocampus_ etc.). We treated each compound as a fixed-effect incorporating the effect size (*r*) and sampling variability (*se*^*2*^_*r*_)for neuronal signatures of a compound. Theoretically, if the transcriptional signature of a compound is negatively associated with a disease, this compound may ‘reverse’ the underlying disease mechanisms and *may* increase the likelihood of demonstrating clinical utility. Hence, we emphasized compounds with negative meta-regression coefficients that also survived correction for multiple testing (FDR < 0.05) and defined these compounds as *potential* therapeutics.

We then used a computational follow-up approach to prioritize the potential therapeutics identified from our drug discovery analysis using two independent transcriptome-wide datasets. These datasets included a human neuronal cocaine exposure model^19^ (SH-SY5Y neuroblastoma cells^;^ GSE71939) and a mouse model of cocaine self-administration^20^ (GSE110344). The neuronal cocaine exposure dataset included 18 samples that assessed mRNA expression (via microarray) for two time points (6 or 24 hours post exposure) across three doses (0*µ*M, 1*µ*M and 5*µ*M; the latter two mimic biologically relevant cocaine levels found in individuals with cocaine abuse). The mouse RNA-sequencing (RNA-seq) data were collected from 12-15 male C57BL/6J mice using similar brain regions to the human samples (ventral tegmental area (VTA), hippocampus and PFC). Mice pressed a lever to receive intravenous infusions of cocaine (1mg/kg; 2 hour sessions for 2 weeks) or saline. Brain tissue was extracted 24 hours after the last self-administration session and mouse modeling was limited to orthologous genes listed from the mouse genome informatics dataset (http://www.informatics.jax.org/).

To maintain consistency, we harmonized the data processing and analyses for our validation datasets to resemble the human brain data (see Supplementary Methods). Using multi-level meta-regression, we assessed whether the potential CUD treatments were also negatively associated with the differential expression of the CUD genes in the neuronal cocaine exposure and mouse cocaine self-administration datasets – accounting for dose and time (neuronal exposure data) or brain region (mouse data) as random effects. Results from the computational follow-up were used to prioritize medications for experimental validation.

### Behavioral assays

*Drosophila melanogaster* (Canton-S B strain) were reared on cornmeal-molasses-yeast medium at 25°C and 70% humidity under a 12h light-dark cycle (lights on at 6:00 am). Cocaine-HCl was obtained from the National Institute on Drug Abuse under Drug Enforcement Administration license RA0443159. Flies were food-deprived for 24 hours followed by a 20 minute free-feeding period. During this period, flies consumed one of three food formulations containing either no drugs (n = 66 females, n = 61 males), cocaine only (n = 141 females, n = 140 males), or cocaine and Ibrutinib (n = 142 females, n = 132 males). The concentrations of cocaine-HCl and Ibrutinib (Tocris Bioscience; Bristol, UK; Product No: 6813; Batch No: 2A/247900) were both 0.16% (w/w). Immediately following exposure, each fly underwent behavioral testing for startle response induced by a mechanical disturbance. A vial housing a single fly was dropped through a chute from a height of 42 cm and then secured in a horizontal position. During the next 45 seconds, the total time spent moving and the occurrence of seizures were recorded. Seizure activity was defined as significant disruption of normal movement with severe tremors and muscle twitching (see Supplementary File S3). While immobile, seizing, or grooming, flies were considered stationary. Startle response data were analyzed using a two way fixed effects factorial ANOVA model: *Y* = *M*+ *S* + *T*+ *S*× *T*+ *ε*, where *Y* is the phenotype, *M* is the mean, *S* is sex, *T* is treatment, *S*×*T* is the sex by treatment interaction, and *ε* is the residual. We used the type III Sums of Squares due to differing sample sizes across groups. Post-hoc comparisons were performed using Student’s *t*-tests. Seizure data were analyzed using Fisher’s exact tests in R.

## RESULTS

### Drug Discovery Input

A schematic overview of the study design is presented in **Figure 1**. Prior to our drug discovery analysis we examined the input data. We observed minimal overlap among differentially expressed genes identified across human brain regions (see Supplementary Figure 1). Associations among KEGG cocaine addiction genes were modest and sometimes negative (*r*_*midbrain_hippocampus*_ = −0.31, *p* = 0.044; *r*_*dlPFC_hippocampus*_ = 0.02, *p* = 0.915; and *r*_*midbrain_dlPFC*_ = 0.36, *p* = 0.019). This heterogeneity and individual variation may be one factor contributing to the difficulty in finding an effective treatment that targets multiple brain regions and is effective for all individuals in a population.

### Drug-Discovery Analyses

Our drug discovery analysis discovered 16 medications with gene expression signatures that were negatively associated with gene expression associated with CUD in the midbrain, hippocampus and dlPFC (*p*_adj_ < 0.05; **Figure 2A**), with diverse pharmacological mechanisms of action (Supplementary Table S2). These medications outperformed the current pharmaceuticals undergoing clinical trials for CUD (**Figure 2B**). Current clinical trial treatments for CUD exhibited more heterogeneity across brain regions and samples than the repurposable medications we identified.

**Figure 2:**
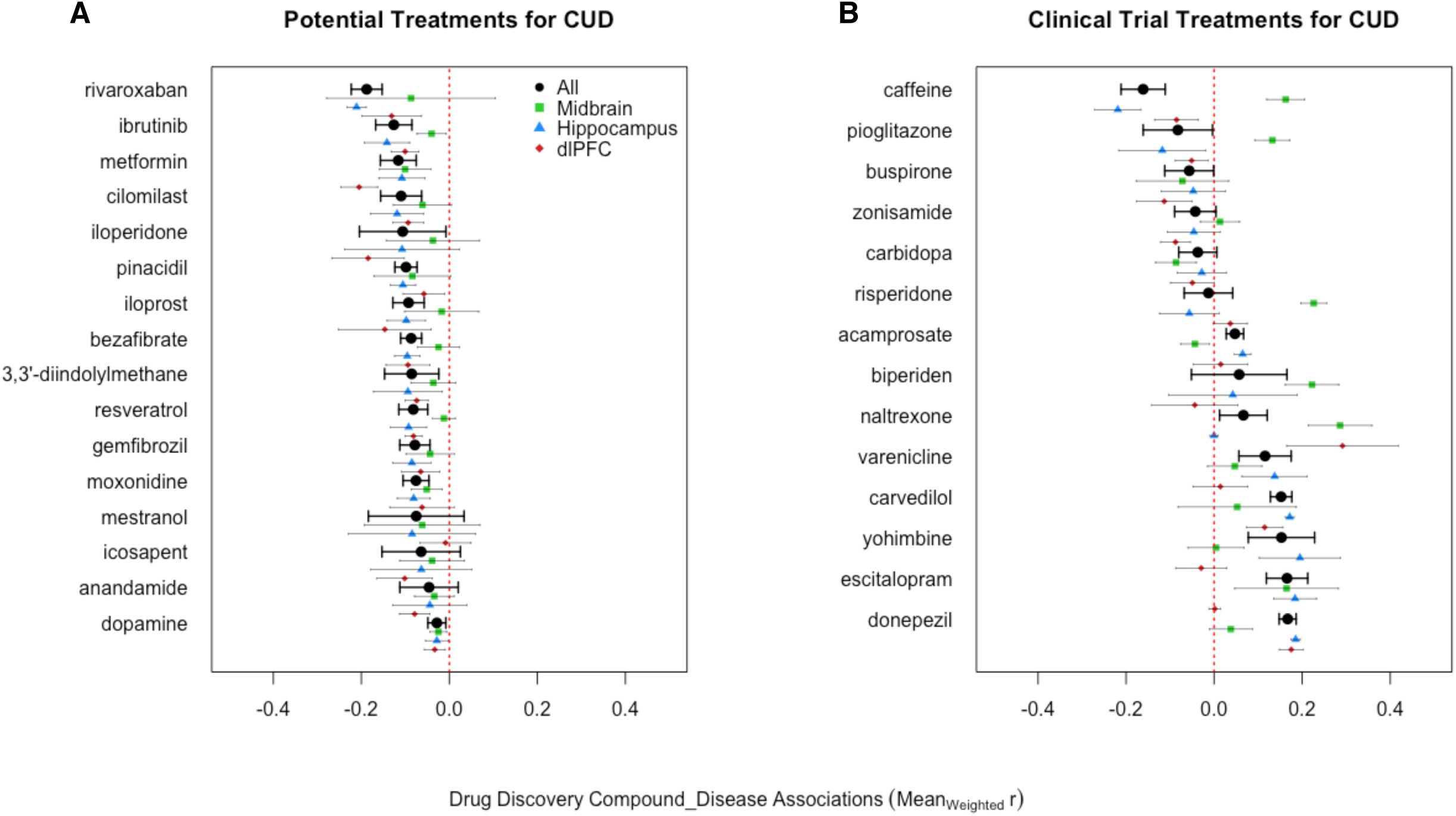
Drug Discovery Results for CUD using Human Brain Data. **A)** shows repurposable compounds negatively associated with CUD gene expression in the brain (all *p*_adj_ < 0.05). **B)** shows the results of the medications currently undergoing clinical trials for CUD. The x-axis represents the weighted average of a compound’s correlation coefficients with the human brain CUD genes and each compound’s weighted standard error bars. Also, note that the black circles represent the effect across all brain regions/studies and the points below are color coded by brain region.

Next, we employed a computational follow-up of the 16 potential CUD medications using transcriptome-wide data from preclinical *in vitro* and *in vivo* models. Only one compound, *Ibrutinib*, was negatively associated with neuronal cocaine exposure (*p* = 0.001; *p*_adj_ = 0.019) and was also trending to reverse gene expression profiles associated with mouse cocaine self-administration (*p* = 0.007; *p*_adj_ = 0.0732; **Figure 3**). No other medication demonstrated significant evidence among preclinical models (see Supplementary Figure S2). Using correlations between human CUD brain data and the *in vitro* drug discovery data, we found that Ibrutinib may reverse the expression of genes involved in dopaminergic and glutamatergic neurotransmission as well as various immediate early genes, intracellular signaling cascade genes and putative neuroepigenetic transcripts (**Figure 4**).

**Figure 3:**
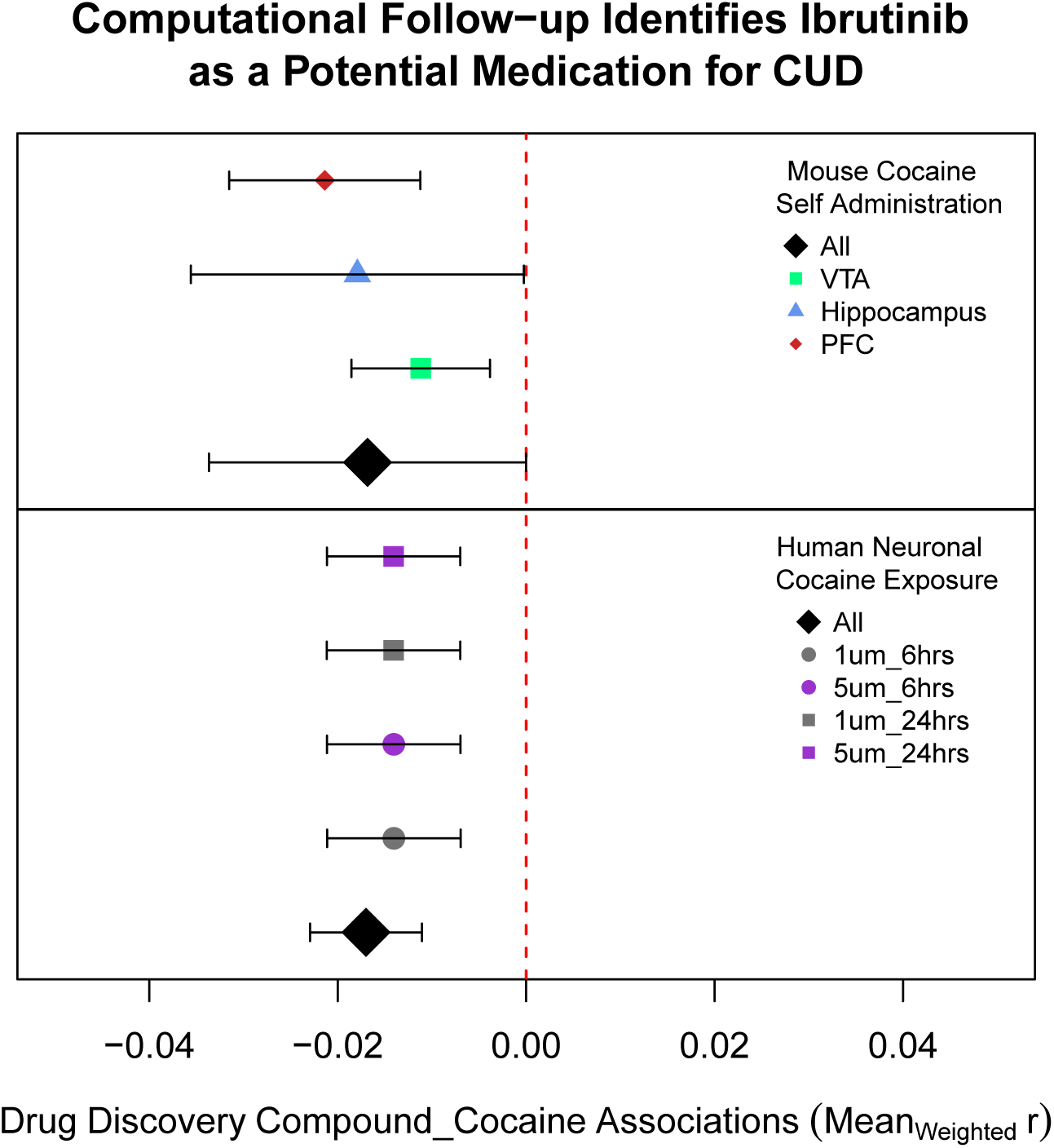
Computational Follow-up in Preclinical Models of Cocaine Use. The bottom panel shows associations between Ibrutinib’s neuronal signatures and diffferential expression results from the *in vitro* cocaine neuronal exposure data. The top panel displays the associations between Ibrutinib’s neuronal signatures with differential expressed genes from the mouse cocaine self-administration dataset.

**Figure 4:**
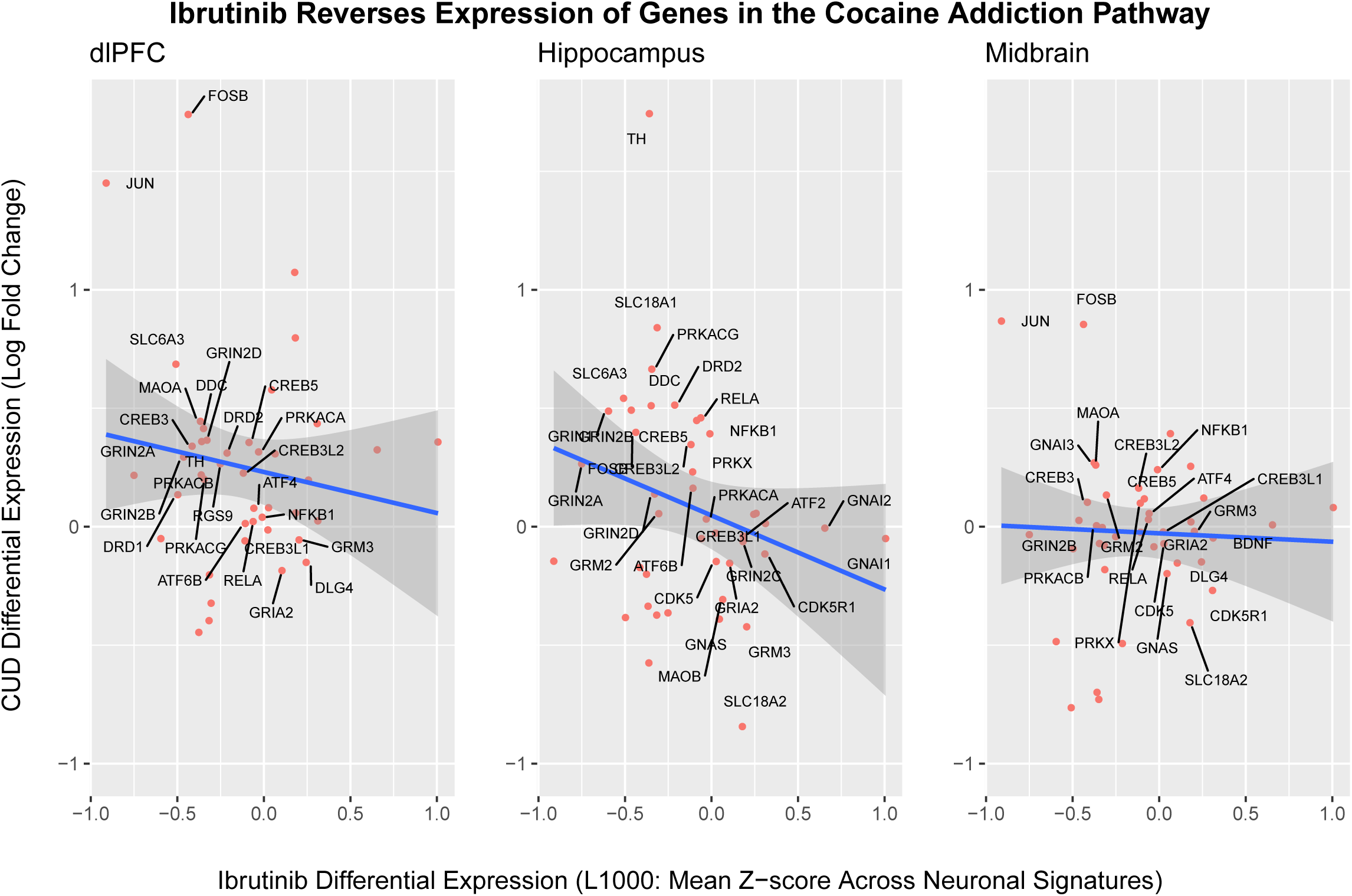
Ibrutinib’s Neuronal Signatures are Negatively Associated with Cocaine Addiction Genes in the Human Brain. Blue line shows the best fitting line and standard error between CUD gene expression (of KEGG Cocaine Addiction Pathway genes) and neuronal signatures of Ibrutinib (collapsing across dose, time and cell line). Genes were labeled if they were in the negative diagonal of the plot. Differential expression of *FOSB* in the dlPFC was windsorized for visualization purposes.

### Validation of potential therapeutic efficacy of Ibrutinib in the Drosophila model

To validate the effectiveness of Ibrutinib as a potential therapeutic for CUD, we evaluated its effects on cocaine-induced phenotypes in the *Drosophila melanogaster* model system. Drosophila provides an advantageous model system for studies on cocaine consumption^21^. The Drosophila dopamine transporter contains a binding site that can accommodate cocaine^22^, and exposure to cocaine gives rise to motor responses that resemble behaviors observed in rodents. In addition, flies develop sensitization to repeated intermittent exposure to cocaine^23,24^.

We measured startle behavior and the prevalence of seizure activity in male and female flies following acute consumption of solid food, solid food supplemented with cocaine, or solid food supplemented with cocaine and Ibrutinib. Male flies exposed to cocaine spent less time moving after being subjected to a mechanical startle (**Figure 5A**). This is likely due to the occurrence of cocaine-induced seizures, which were scored as stationary periods. Both male and female flies that consumed Ibrutinib with cocaine spent more time moving than flies that only consumed cocaine (**Figure 5A**). Male flies that consumed Ibrutinib and cocaine showed a significant decrease in the prevalence of seizures (*p* = 0.0029; **Figure 5B**). Fewer cocaine-induced seizures were also observed in Ibrutinib treated females, but this observation did not reach statistical significance since the incidence of cocaine-induced seizures was lower in females than in males. These results confirm that Ibrutinib may be useful as a therapeutic to prevent various neurobehavioral and neurotoxic effects of cocaine use.

**Figure 5:**
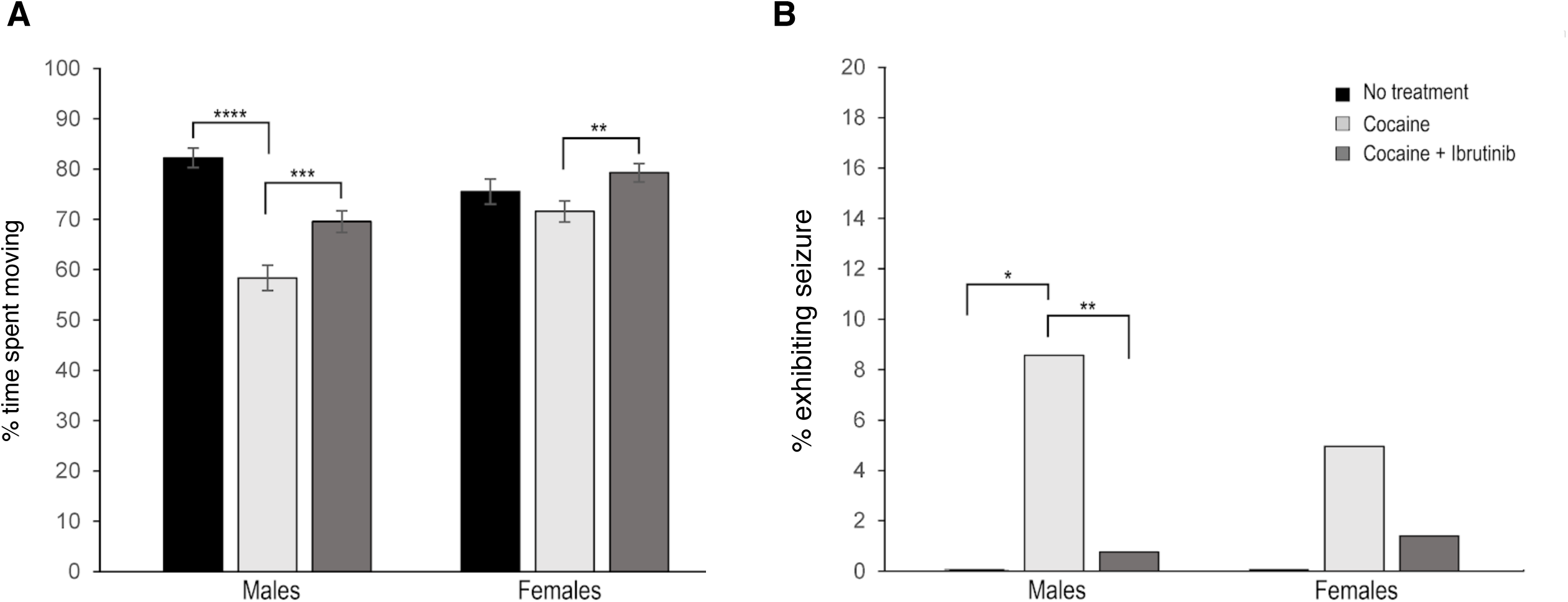
Ibrutinib Mitigates Cocaine-induced Startle Response and Cocaine-induced Seizures in *Drosophila melanogaster*. Sample sizes were as follows: (n = 61 (♂control), 140 (♂cocaine), 132 (♂cocaine + Ibrutinib), 66 (♀control), 141 (♀cocaine), 142 (♀cocaine + Ibrutinib) **(A)** Startle response. The percent time out of 45 seconds that each fly spent moving following a 42 cm drop was measured. Male flies exposed to cocaine spent less time moving than flies exposed to untreated food (*p* = 2.13 x 10^−12^). Male and female flies exposed to cocaine and Ibrutinib exhibited an increase in the percent of time spent moving compared to flies only exposed to cocaine (♂ *P* = 0.000824, ♀ *P* = 0.00246; two-tailed Student’s t-test). Error bars represent standard error. **(B)** Seizure activity during the startle response. The number of flies that exhibited seizure activity during the startle assay was recorded. Male flies exposed to cocaine exhibited more seizure activity than flies exposed to untreated food (*p* = 0.0195; Fisher’s exact test). Male flies exposed to cocaine and Ibrutinib exhibited a decrease in the frequency of seizures compared to flies only exposed to cocaine (*p* = 0.0029; Fisher’s exact test).

## Discussion

We used a novel technique to identify and validate potential repurposable medicaitons for CUD or cocaine toxicity. These medications demonstrated more reproducible associations across the brain’s reward circuitry than the current medications undergoing clinical trials in the US. Despite the low correlation of gene expression across datasets, Ibrutinib consistently reversed patterns of gene expression observed in brains of individuals with CUD across brain regions and diverse samples as well as across two gold standard preclinical models. Thus, we consider Ibrutinib to be the most promising repurposable medication for CUD identified by our study.

Ibrutinib is a Bruton’s tyrosine kinase inhibitor and is approved by the FDA for various B cell cancers including chronic lymphocytic leukemia^25^. While Ibrutinib is not a typical candidate for addiction therapeutics, our study suggests that it may target and reverse the expression of cocaine addiction genes in human meso-cortico-limbic circuitry. Notably, Ibrutinib shares a pharmacological mechanism of action with Terric acid, a compound that reduced binge drinking in mice^13^.

Ibrutinib may have therapeutic potential for cocaine neurotoxicity. Our results from the Drosophila models suggest Ibrutinib significantly reduces cocaine-induced seizure frequency. While rare, cocaine-induced seizures have a serious and urgent need for treatments^26^. The precise mechanism by which Ibrutinib may alleviate cocaine neurotoxicity remains unclear. Ibrutinib influences intracellular cascades involved in pro-inflammatory response, Ca^+^ signaling and protein kinase activity^27, 28^. These processes are also involved in cocaine use^29, 30^, and cocaine-induced seizures^31^.

Most therapeutics for substance use disorders (and drug-induced overdoses) target neurotransmitter receptors. We found that expression of the dopamine transporter (*SLC6A3* or *DAT1*) as well as various dopamine and glutamate receptors (*DRD1, DRD2, GRIN2D, GRIN2A, GRIN2B, GRIA2*) in brains from CUD individuals were negatively associated with the *in vitro* neurotranscriptional signatures of Ibrutinib. These findings are correlative, but illuminate possible pleiotropic mechanisms in which Ibrutinib counteracts the neuropathology of CUD. Finally, both cocaine and Ibrutinib act on the mushroom bodies in the fly brain. These central brain structures are responsible for associative learning and memory^32^, and experience-dependent behavioral modification^32^. The mushroom bodies also serve as initiation sites for seizures^33^ and drug consumption^34^.

In addition to Ibrutinib, we discovered other potential medications that may counter some of the neuroadaptations of CUD. Some of these compounds may target molecular processes underlying chronic cocaine use. For instance, we found that dopamine reversed the differential expression of CUD genes across brain regions, which may counteract the hypodopaminergic state induced by chronic cocaine use^35^. A preponderance of the drugs tested seemed to be sensitive to various cardiac complications associated with cocaine use, such as Rivaroxaban, Pinacidil, Iloprost, Gemfibrozil, Bexafibrate and Moxonidine. It is possible that these medications exert a neuroprotective role for cocaine use and may be relevant for cocaine toxicity. Some of these medications may have both health and behavioral benefits. For instance, Moxonidine reduces (cue-induced) cocaine relapse^36^ and ethanol withdrawal^37, 38^ in rodents. Of note, the pharmacological profiles of the potential CUD medications we identified have shown to reduce various drug use behaviors (PPAR alpha agonists^39^; PDE4 inhibitors^40,41^). Potential CUD medications also influenced catecholamine neurotransmission, potassium channels, hormonal signaling and mitochondrial processes, which have all been implicated in the pathophysiology of cocaine use^42,43, 44, 45^. Our results may serve a dual purpose for indirectly unraveling neuromolecular features of human CUD while also identifying specific chemical compounds that may demonstrate clinical utility for cocaine-related outcomes.

The results of our study should be interpreted with caution until further validation studies are conducted. We used all extant transcriptome-wide brain data on human CUD, but these sample sizes were small and were ascertained from highly selected cocaine users. Additionally, these data are unable to characterize specific symptoms of CUD and may have bias relating to health and psychiatric comorbidities. We sought to minimize these biases by investigating validated genes from the cocaine addiction (KEGG) pathway as well as integrating tightly controlled experimental cocaine data from preclinical paradigms. Our study benchmarked potential CUD medications against current targets in clinical trials, although we did not have data on various pharmaceuticals that have demonstrated some efficacy for individuals with CUD (e.g., disulfiram^46^; topiramate^47^). The lead repurposable drug from this study, Ibrutinib, may not be an accessible treatment option to many, as it can be costly^48^ and may be associated with side effects^49^. In addition, it is unknown what dose would be most effective and whether negative health consequences arise when combining this medication with cocaine (or crack cocaine). As with other substance use disorders, pharmaceutical interventions for CUD will likely only provide incremental improvement. The most successful treatment will likely require a combination of medication and psychosocial interventions such as cognitive-behavioral therapy, contingency management, and lifestyle modification as a form of relapse prevention^50^. Despite these limitations, Ibrutinib emerges as a promising therapeutic with potential for the treatment of CUD-associated symptoms. In a broader context, our study provides a proof-of-principle for a powerful, flexible and interpretable method that can be used to identify and prioritize therapeutics from genome-wide and transcriptome-wide datasets.

## Data Availability

Data used for drug discovery analyses are publicly available on Gene Expression Ominbus, Sequence Read Archive and Array Express.

## Acknowledgements

We acknowledge the National Institute on Drug Abuse award DP1DA042103 (awarded to RHCP) and DA041613 (awarded to RRHA and TFCM). We are grateful for the LINCs database curators as well as those responsible for the availability of the RNA-sequencing and microarray data. Content is solely the responsibility of the authors and does not necessarily represent the official views of the National Institutes of Health or the Department of Veteran Affairs.

## Conflict of Interest

The authors declare no conflicts of interest.

## Notes

### Competing Interest Statement

The authors have declared no competing interest.

### Author Declarations

Data used in this study is exempt from IRB oversight.

