## Supplementary Information for "Ibrutinib as a Potential Therapeutic for Cocaine Use Disorder"

*Supplementary Methods*

*Data Processing and Analysis*

We analyzed data from two microarray studies: human midbrain samples^1^ and *in vitro* neuronal cocaine exposure data^2^. Human midbrain samples contained three technical replicates for each individual. Correlations across technical replicates were high (all r > 0.95). Thus, we averaged across individual samples before normalization, which mitigates potential biases of batch effects. The *in vitro* neuronal cocaine exposure data only contained biological replicates, but batch effects were controlled for using surrogate variable analyses^3^. All microarray samples contained multiple array ids for individual genes. To obtain a single expression value for a gene we used the *collapseRows* function that refined all array ids into feature intensity recordings for specific genes. All genes’ feature intensity recordings were variance stabilized (via *vsn2* command^4^) and then loess normalized (via the *normalizeCyclicLoess* command^5^) before differential expression analysis. Our study used a linear empirical Bayes to obtain differential expressed genes for microarray data via the *eBayes* command^5^.

Our study also utilized three separate RNA-sequencing (RNA-seq) samples: human dorsal-lateral prefrontal cortex^6^ (dlPFC), human hippocampus^7^ and mouse self-administration data^8^ (PFC, hippocampus and ventral tegmental area (VTA)). We used standard DESeq2^9^ practices to estimate differentially expressed genes for RNA-seq data. Briefly, all data was normalized with the *estimateSizeFactors* command that performs a standard median ratio method. Samples with that were < 2 standard deviations from the mean normalization value were considered outliers and removed (one sample from the human hippocampus). Subsequently, dispersion was adjusted for via the *estimateSizeFactors* command that performs a Cox Reid-adjusted profile likelihood maximization function. Controlling for batch effects (via svaseq^10^; 2 surrogate variables), we then used the *nbinomWaldTest* command that leverages a generalized linear model and assumes a negative binomial distribution to estimate differential expression. Note, that the differentially expressed genes estimated from our study were highly correlated with the originally published work (all *r* > 0.95).

| **Post-Mortem Human Sample Information: M (s.d.) [n]** | | | |
| --- | --- | --- | --- |
| Attribute | dlPFC [n = 36] | Hippocampus [n = 15] | Midbrain [n = 20] |
| Specific Brain | Broadmann's Area 46 | CA4-CA1 and | VTA & SN |
| Region |  | Dentate Gyrus |  |
| CUD | DSM-V | DSM-IV Abuse | DSM-IV Abuse |
| Criteria |  | or Dependence |  |
| Technology | RNA-seq | RNA-seq | Microarray |
| n [case / control] | [19 / 17] | [7 / 8] | [10 / 10] |
| Sex | 100% Male | 100% Male | 100% Male |
| Age | 35.0 (11.0) | 39.4 (6.4) | 49.2 (3.9) |
| Ethnicity | European American or Hispanic [23] | European American or Hispanic [10] | European American or Hispanic [4] |
|  | African American [13] | African American [5] | African American [16] |
| PMI | 16.5 (6.4) | 16.9 (4.2) | N/A |
| Brain pH | 6.4 (0.3) | N/A | 6.5 (0.2) |
| RNA Integrity | 2.9 (0.8) | N/A | 6.5 (0.9) |

**Supplementary Table S1:** Descriptive Statistics on Brain Tissues and Samples

Note the incomplete information across samples (e.g., missing PMI [post-mortem interval], RNA integrity, incomplete information on ethnicity and other aspects).

**Supplementary Figure S1** Negligible Overlap of Differentially Expressed Genes Associated with CUD (FDR < 0.05) Across Brain Regions / Study

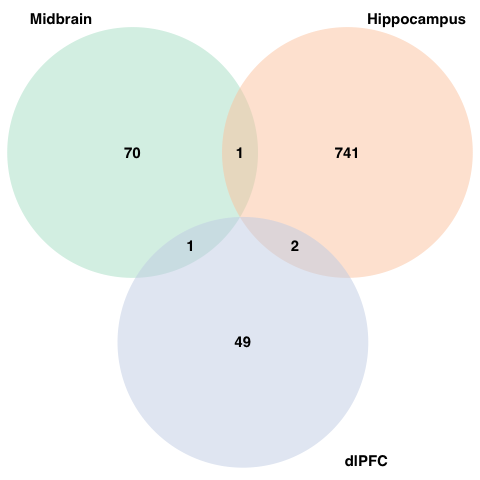

Note these are the # of genes per each brain region that were also included in the L1000 drug discovery dataset.

**Supplementary Table S2** Descriptive Information on the Potential Therapeutics Identified for CUD

| **Proposed Mechanisms and Gene Targets for the Potential Repurposable Medications for CUD or Cocaine Toxicity** | | |
| --- | --- | --- |
| Compound | Mechanism of Action | Gene Targets |
| **ibrutinib** | Bruton's tyrosine kinase (BTK) inhibitor | *BLK, BMX, BTK* |
| rivaroxaban | coagulation factor inhibitor | *F10* |
| pinacidil | ATP channel activator & | *ABCC8, ABCC9* |
|  | potassium channel activator |  |
| iloperidone | dopamine receptor antagonist & | *ADRA1A, ADRA2C, DRD1, DRD2, DRD3, DRD4,* |
|  | serotonin receptor antagonist | *HRH1, HTR1A, HTR2A, HTR6, HTR7* |
| metformin | insulin sensitizer | *ACACB, PRKAB1* |
| anadamide | cannabinoid receptor agonist | *CACNA1G, CACNA1H, CACNA1I, CNR1, CNR2* |
|  |  | *GLRA1, GPR18, GPR55, KCNA2, KCNK3, KCNK9, TRPM8, TRPV1* |
| icosapent | platelet aggregation inhibitor | *ACSL3, ACSL4, FADS1, FFAR1, PPARD,* |
|  |  | *PPARG, PTGS1, PTGS2, SLC8A1, TRPV1* |
| dopamine | dopamine receptor agonist | *DBH, DRD1, DRD2, DRD3, DRD4, DRD5,* |
|  |  | *HTR1A, HTR7, SLC6A2, SLC6A3, SLC6A4* |
| iloprost | platelet aggregation inhibitor & | *PTGDR, PTGER1, PTGER2, PTGER3,* |
|  | prostanoid receptor agonist | *PTGER4, PTGFR, PTGIR, TBXA2R* |
| cilomilast | phosphodiesterase inhibitor | *PDE4A, PDE4B, PDE4D* |
| reservatrol | cytochrome P450 inhibitor, SIRT activator | *CSNK2A1, NQO2, PTGS1, PTGS2* |
| mestranol | estrogen receptor agonist | *ESR1* |
| gemfibrozil | lipoprotein lipase activator | *LPL, PPARA, SLCO1B1, SLCO1B3, SLCO2B1* |
| bezafibrate | PPAR receptor agonist | *PPARA, PPARD, PPARG* |
| moxonidine | imidazoline receptor agonist | *ADRA2A, ADRA2B, ADRA2C* |
| 3,3' - diindolylmethane | CHK inhibitor, cytochrome P450 activator & | *AR, HIF1A, IFNG, PI3* |
|  | indoleamine 2,3-dioxygenase inhibitor |  |
| rivaroxaban | coagulation factor inhibitor | *F10* |

**Supplementary Figure S2** Computational Follow-up of Potential Therapeutics for CUD in Preclinical Models of Cocaine Use

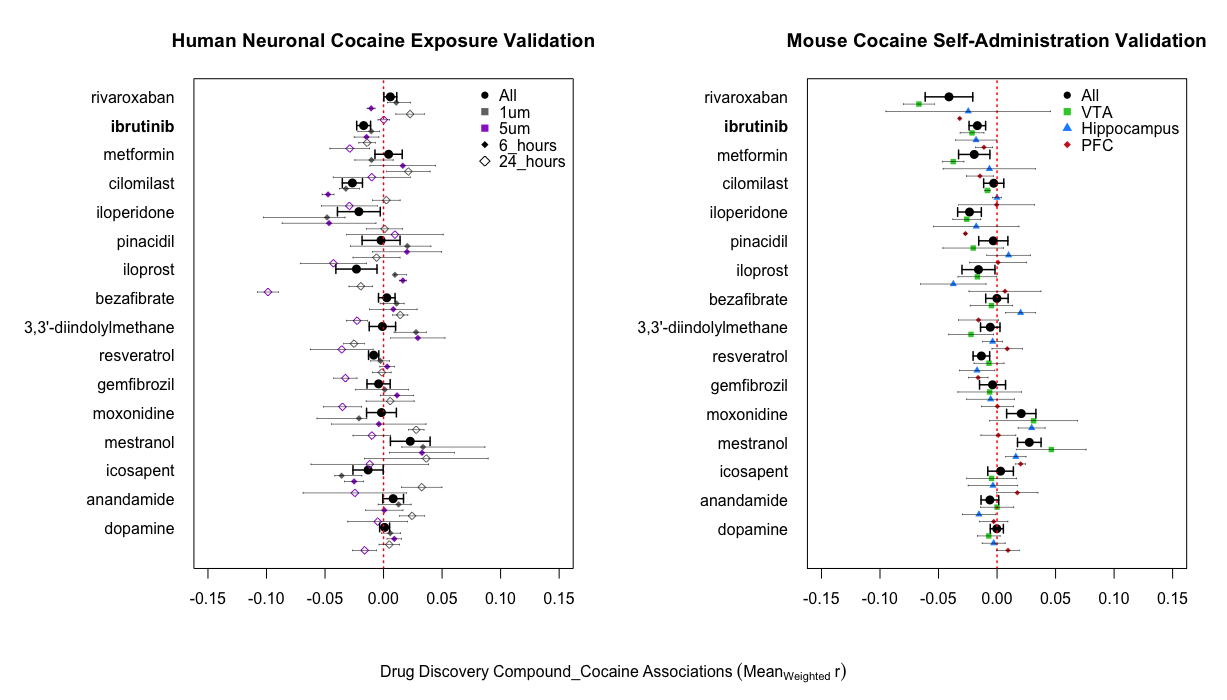

The left panel shows the results from the *in vitro* cocaine exposure dataset^2^ whereas the right panel displays findings from the *in vivo* dataset^8^. *Note* the points on the graph are color coded by dose and time (left panel) and brain region (right panel), with the black circle representing the overall effect for a compound.
